# Longitudinal Changes in Anatomic Biomarkers on Optical Coherence Tomography Angiography in Diabetic Retinopathy

**DOI:** 10.1101/2025.08.07.25333210

**Authors:** Rachel Liu, Isaac Bakis, Megan Steinkerchner, Kevin Sun, Sapna Gangaputra, Stephen Kim, Lok Hin Lee

**Affiliations:** Vanderbilt University School of Medicine; Vanderbilt Eye Institute

## Abstract

**Purpose:** To identify optical coherence tomography angiography (OCTA)-derived biomarkers correlating with diabetic retinopathy (DR) severity and characterize longitudinal retinal microvascular changes across DR stages.

**Methods:** In this 3-year prospective study, we analyzed OCTA images from 328 eyes of 164 adults with type II diabetes and 33 eyes from 17 healthy controls. Patients were categorized as no DR, mild, moderate, or severe nonproliferative DR (NPDR), or proliferative DR (PDR) at baseline. Bilateral OCTA scans were obtained at each visit and processed with a validated pipeline to extract seven microvascular indices from the superficial and deep capillary plexuses: vessel density, skeleton density, acircularity index, average vessel caliber, foveal avascular zone (FAZ) area, FAZ perimeter, and fractal dimensions. Linear regression quantified changes over time, and intergroup comparisons were made using ANOVA and post-hoc testing.

**Results:** Patients with severe NPDR showed a significant annual decrease in vessel density and skeleton density in the deep capillary plexus compared to healthy controls, indicating progressive ischemia. Acircularity index increased significantly in severe NPDR compared to mild NPDR, suggesting worsening macular ischemia. In the superficial plexus, severe NPDR patients exhibited significantly greater annual vessel caliber narrowing than mild NPDR. Conversely, skeleton density in PDR increased relative to severe NPDR, possibly reflecting neovascular remodeling.

**Conclusions:** Longitudinal OCTA analysis reveals stage-specific microvascular changes in DR. These dynamic biomarkers may provide early indicators of DR progression and offer a quantitative foundation for individualized monitoring and timely intervention strategies.

**Translational Relevance:** OCTA-derived biomarkers offer a noninvasive tool for detecting early DR changes and guiding personalized care strategies.

## Introduction

Diabetic retinopathy (DR) is characterized by microvascular ischemia leading to progressive damage to the retinal microvasculature and subsequent retinal neurodegeneration [1]. More recently, optical coherence tomography angiography (OCTA) has emerged as an alternative to invasive and time-intensive fundus fluorescein angiogram (FFA) and provides high-resolution images of both the deep and superficial capillary plexuses of the retina [2,3].

Several studies have demonstrated the utility of OCTA in characterizing the severity of DR using microvascular parameters. Lee et al. demonstrated that vessel tortuosity increased with the severity of non-proliferative DR (NPDR) but decreased in proliferative DR (PDR), suggesting alterations in vascular remodeling during the progression of DR [4]. Sun et al. revealed that OCTA parameters like foveal avascular zone (FAZ) area, vessel density, and fractal dimension were significantly associated with DR progression and the development of diabetic macular edema (DME) [5].

Despite these promising findings, there are limitations in the current body of literature. Most studies on OCTA and DR have been cross-sectional, providing only a snapshot of microvascular changes at a single point in time [4, 6]. While these studies have established correlations between OCTA parameters and DR severity, they are unable to provide insight into how these parameters change longitudinally as the disease progresses or regresses in response to treatment. Current longitudinal studies on OCTA and DR are limited in breadth and scope, often including only one stage of DR or only one microvasculature metric [5, 7]. Additionally, there is a lack of standardization in OCTA acquisition protocols and quantitative metrics across studies, making direct comparisons difficult.

Our study aims to longitudinally assess changes in retinal microvascular indices on OCTA across different stages of DR. We aim to characterize OCTA biomarkers that correlate with DR severity. This will improve the clinical utility of OCTA in monitoring the changes in DR over time and detecting pre-clinical changes in retinal microvasculature.

## Materials and Methods

### Study design and population

This study utilized OCTA images collected during the Inflammatory Mediators in the Pathophysiology of Diabetic Retinopathy (INSPIRE) trial, a single-center prospective 3-year randomized clinical trial conducted at Vanderbilt Eye Institute which enrolled 328 eyes of 164 adult patients with type II diabetes [8]. Participants were initially categorized into no DR, NPDR, and PDR based on the International Clinical Disease Severity Scale. Patients in the NPDR cohort were further divided into mild NPDR, moderate NPDR, and severe NPDR based on clinician categorization for image analysis. Participants underwent bilateral OCTA imaging every 4 months for 3 years, with 5 repeat scans of the same eye at each visit. 33 eyes from 17 healthy control patients underwent yearly OCTA imaging. The study adhered to ethical guidelines, receiving approval from the Vanderbilt University Medical Center Institutional Review Board (#201433), and all patients provided written informed consent. It complied with HIPAA regulations and the Declaration of Helsinki, with trial registration at ClinicalTrials.gov (NCT04505566) and funding from the National Eye Institute (R01-EY031315).

### Analysis of optical coherence tomography angiography

Retinal vessel metrics were extracted using a systematic image-processing pipeline applied to en-face OCT images that has previously been reported and validated [9–11]. Each image underwent a top-hat morphological filtering step to enhance vessel contrast and diminish background intensity fluctuations. This was followed by a Hessian-based filter designed to highlight tubular structures by emphasizing their second-order intensity patterns. To create a preliminary vessel map, the resulting output was subjected to Huang’s entropy-based thresholding, which provides an adaptive global threshold. In parallel, a local median adaptive thresholding procedure was independently conducted on the preprocessed OCTA images. The two threshold images were then merged by selecting the pixels common to both, to create a vessel segmentation map. Finally, the resulting vessel map was skeletonized, reducing each vessel to a single-pixel-wide representation suitable for downstream quantitative analyses.

FAZ segmentation was performed using a morphological Chan-Vese (MCV) active contour approach [10–15]. The original OCTA images were first smoothed with a Gaussian filter to reduce high-frequency noise and improve edge definition. A circular initial level set was positioned near the geometric center of each image to guide the MCV model’s iterative evolution toward the anatomical location of the FAZ. Following convergence, a binary mask of the FAZ was generated and further refined by filling any small gaps or artifacts. To ensure measurement accuracy, only the largest contiguous region within the mask was preserved, delineating the FAZ boundary for subsequent quantification of FAZ metrics, including area, perimeter, and acircularity index.

From the vessel segmentation and FAZ boundary, seven microvasculature indices were extracted from the deep and superficial layers of the OCTA images: acircularity index, average vessel caliber, FAZ area, FAZ perimeter, fractal dimensions, skeleton density, and vessel density (Figure 1). Definitions of these indices are listed below, and representative images of selected indices are shown in Figure 2.

1. **Acircularity index**: A dimensionless measure of irregularity and ischemia in the avascular region of the central fovea
2. **Average vessel caliber (um)**: The average diameter of all vessels in scanned area
3. **FAZ area (mm^2^)**: An area measurement of the central region of the fovea that lacks capillaries
4. **FAZ perimeter (mm)**: A measurement of the central fovea’s FAZ perimeter
5. **Fractal dimension**: A mathematical measurement of the branching of retinal vasculature
6. **Skeleton density (%)**: The percentage of the total retinal area covered by vessels, where each vessel is converted into a skeletonized line, and the total line length is calculated as a percentage of the retinal area.
7. **Vessel density (%)**: The percentage of the total retinal area that is covered by vessels.

**Figure 1.**
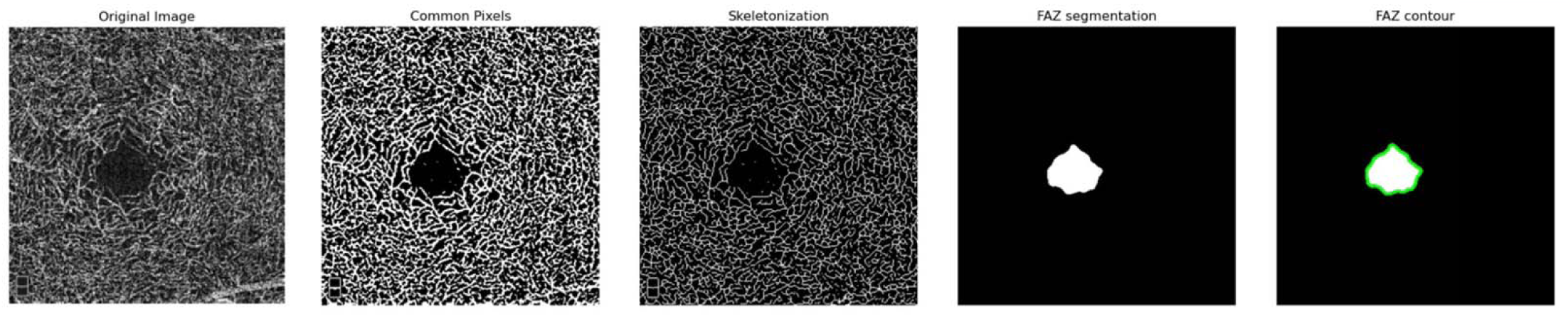
Post-segmented images for metric extraction. Common pixels were used to calculate vessel density and average vessel caliber. Skeletonized image was used to calculate skeleton density and fractal dimensions. FAZ segmentation was used to calculate FAZ area, perimeter, and acircularity index.

**Figure 2.**
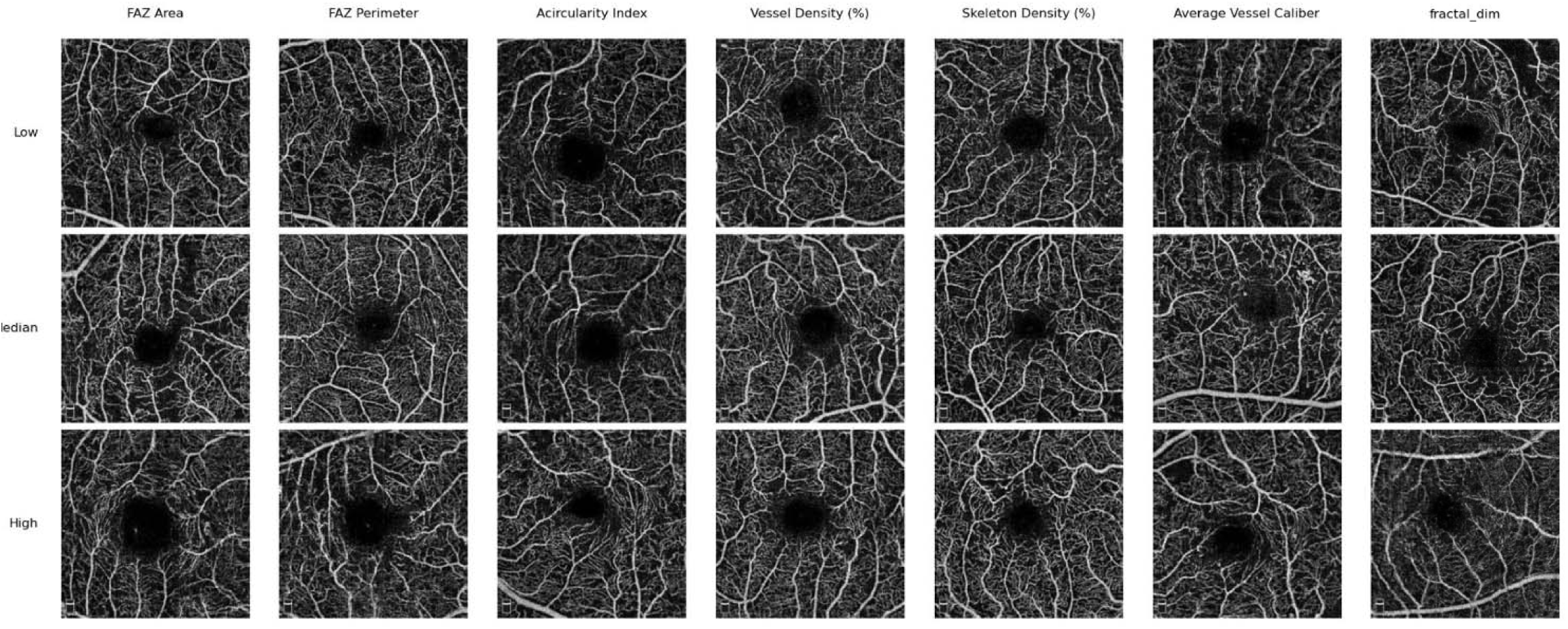
Representative en-face OCT-angiography slabs of the superficial vascular plexus. Columns correspond to seven metrics analyzed in the study. Rows show eyes in the cohorts Low (<20th percentile), Median (50th percentile), and High (>80th percentile) values for the metric in that column. In each column, the eye was selected to minimize Mahalanobis distance from the cohort median across all non-target metrics, ensuring that the primary visual distinction is driven by the illustrated parameter.

### Statistical analysis

Statistical analysis was performed using RStudio software (version 4.4.2). Baseline characteristics of patients in each DR category were compared using one-way analysis of variance (ANOVA). To reduce the variability of quantitative metrics, we screened OCTA images for signal strength prior to statistical analysis, and images with a signal strength of ≤ 5 were excluded from analysis. Representative images showing scans of different signal strengths are shown in Figure 3. For each patient visit, the mean value of each image with signal strength > 5 was used for subsequent analysis, as we find qualitatively that this is sufficient to reduce background noise and improve quantitative analysis [12]. To evaluate the progression of disease in terms of microvascular factors, linear regression analyses were performed to assess changes in each factor over time. One-way ANOVA compared the slopes of the linear regressions between patients with different DR categories to evaluate the rate of progression in each factor. Post-hoc Tukey’s tests identified significant (*p* value ≤ 0.05) pairwise intergroup differences in the rate of change of vessel metrics for different classes.

**Figure 3.**
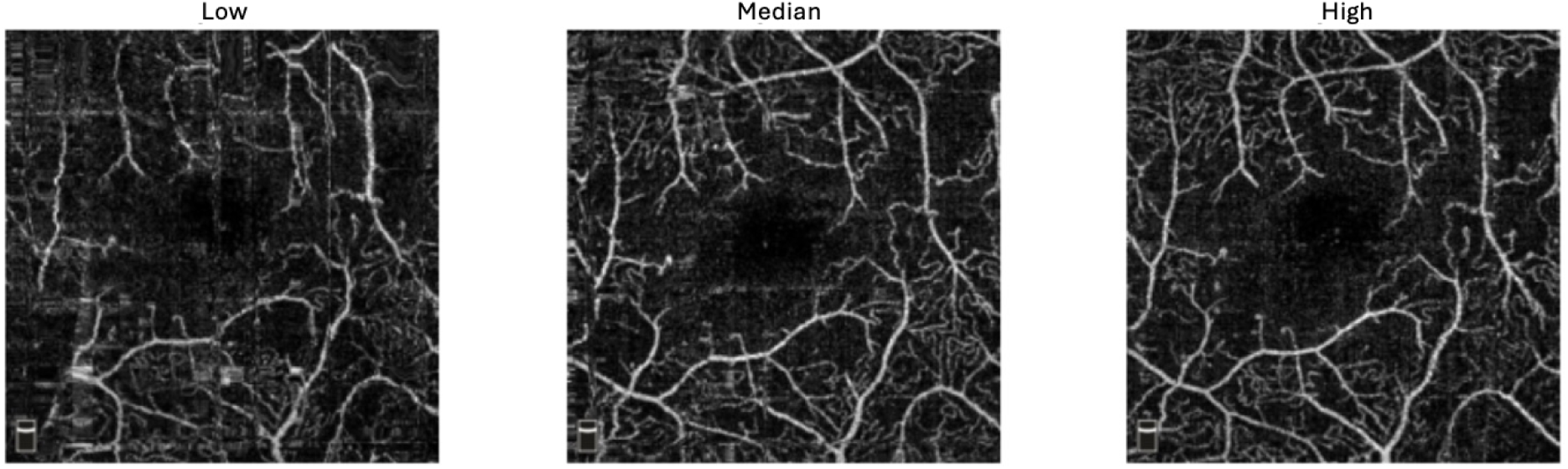
Representative en-face OCT-angiography slabs of the superficial vascular plexus with differing signal strengths. Figure shows images of low (signal strength= 3), median (signal strength = 5) and high-quality (signal strength = 8) images of the same patient. The images were taken within the same hour of each other and variations in quality was due to natural clinical process and not artificially induced.

## Results

A total of 328 eyes from 164 adult patients with type II diabetes and 33 eyes from 17 healthy controls were included in this study. From the collected images, 18% of total scans were excluded due to poor signal strength and images from 7 eyes were excluded for lack of data availability. Images from a total of 55 eyes from patients with diabetes but no diabetic retinopathy (NDR), 7 eyes with mild NPDR, 216 eyes with moderate NPDR, 6 eyes with severe NPDR, 37 eyes with proliferative DR, and 33 eyes from healthy controls were selected for analysis, yielding a total of 321 eyes. Images from healthy controls were obtained every 12 months for 3 years. Baseline demographic data is shown in Table 1, and baseline OCTA quantitative metric values are shown in Table 2.

**Table 1.** Baseline Demographic Characteristics.

|  | *Overall* | *NDR* | *Mild NPDR* | *Moderate NPDR* | *Severe NPDR* | *PDR* | *p value* |
| --- | --- | --- | --- | --- | --- | --- | --- |
| *Number of eyes* | 321 | 55 | 7 | 216 | 6 | 37 | <0.001 |
| *HbA1c, % (Median [IQR])* | 7.5 [6.80-8.60] | 6.50 [6.30-8.10] | 7.65 [7.22-8.03] | 7.60 [6.90-8.65] | 7.90 [7.83-10.53] | 7.70 [6.40-8.90] | 0.076 |
| *CSF Thickness (Median [IQR])* | 282.50 [257.00-313.75] | 277.50 [258.00-295.75] | 299.50 [294.75-329.00] | 282.00 [257.25-314.00] | 310.00 [268.00-313.00] | 267.50 [249.25-323.25] | 0.097 |
| *Macular Volume (Median [IQR])* | 8.58 [8.16-9.14] | 8.36 [8.15-8.69] | 8.56 [8.21-9.14] | 8.64 [8.21-9.16] | 8.58 [8.34-8.95] | 8.29 [8.10-9.17] | 0.299 |
| *Visual Acuity (logMAR, mean ± SD)* | 0.15 ± 0.31 | 0.15 ± 0.19 | 0.04 ± 0.07 | 0.12 ± 0.32 | 0.47 ± 0.86 | 0.18 ± 0.20 | 0.213 |
| *Scans with DME* | 52 (19.94%) | 0 (0%) | 2 (25%) | 34 (18.38%) | 1 (16.67%) | 9 (25.00%) | 0.032 |
| *Average number of visits* | 2.353 | 5.95 | 6.43 | 6.66 | 6.25 | 6.59 | <0.001 |
| *% scans excluded* | 18.4 | 12.1 | 9.3 | 19.3 | 11.4 | 24.7 | <0.001 |

\* NDR: no diabetic retinopathy
NPDR: non-proliferative diabetic retinopathy
PDR: proliferative diabetic retinopathy
HbA1c: hemoglobin A1c
CSF: central subfield thickness
logMAR: logarithm of the minimum angle of resolution
DME: diabetic macular edema

**Table 2:** Baseline OCTA characteristics in the deep and superficial retinal layers. Data presented as mean ± standard deviation.

|  | Healthy controls | NDR | Mild NPDR | Moderate NPDR | Severe NPDR | PDR | p value |
| --- | --- | --- | --- | --- | --- | --- | --- |
| **Superficial Layer** |  |  |  |  |  |  |  |
| *Acircularity* | 1.353 ± 0.160 | 1.552 ± 0.309 | 1.526 ± 0.273 | 1.897 ± 0.493 | 2.377 ± 0.473 | 2.181 ± 0.614 | <0.001 |
| *Average Vessel Caliber* | 22.303 ± 0.487 | 23.038 ± 0.651 | 23.296 ± 0.472 | 23.352 ± 1.115 | 24.172 ± 0.538 | 23.716 ± 1.144 | <0.001 |
| *FAZ Area* | 0.323 ± 0.084 | 0.376 ± 0.140 | 0.312 ± 0.109 | 0.501 ± 0.297 | 0.521 ± 0.169 | 0.623 ± 0.355 | <0.001 |
| *FAZ Perimeter* | 2.721 ± 0.554 | 3.328 ± 1.020 | 2.988 ± 0.803 | 4.718 ± 2.300 | 6.040 ± 1.791 | 3.260 ± 2.181 | <0.001 |
| *Fractal Dimension* | 1.763 ± 0.003 | 1.755 ± 0.008 | 1.754 ± 0.005 | 1.741 ± 0.019 | 1.738 ± 0.009 | 1.731 ± 0.021 | <0.001 |
| *Skeleton Density* | 15.867 ± 0.625 | 14.580 ± 1.095 | 14.207 ± 0.454 | 13.134 ± 1.715 | 12.606 ± 0.843 | 12.137 ± 1.629 | <0.001 |
| *Vessel Density* | 35.839 ± 1.090 | 33.969 ± 1.981 | 33.523 ± 0.841 | 30.924 ± 13.027 | 30.850 ± 1.724 | 29.022 ± 2.946 | <0.001 |
| **Deep Layer** |  |  |  |  |  |  |  |
| *Acircularity* | 1.154 ± 0.0874 | 1.247 ± 0.283 | 1.303 ± 0.311 | 1.481 ± 0.460 | 1.448 ± 0.351 | 1.615 ± 0.482 | <0.001 |
| *Average Vessel Caliber* | 19.906 ± 0.282 | 20.409 ± 0.484 | 20.623 ± 0.431 | 21.189 ± 0.717 | 21.699 ± 0.522 | 21.855 ± 0.725 | <0.001 |
| *FAZ Area* | 0.200 ± 0.068 | 0.223 ± 0.139 | 0.190 ± 0.157 | 0.304 ± 0.237 | 0.261 ± 0.142 | 0.375 ± 0.280 | <0.001 |
| *FAZ Perimeter* | 1.803 ± 0.348 | 2.038 ± 1.312 | 1.936 ± 1.414 | 2.796 ± 1.868 | 2.574 ± 1.297 | 43.419 ± 2.004 | <0.001 |
| *Fractal Dimensions* | 1.776 ± 0.002 | 1.773 ± 0.004 | 1.772 ± 0.004 | 1.766 ± 0.008 | 1.764 ± 0.006 | 1.759 ± 0.009 | <0.001 |
| *Skeleton Density* | 18.430 ± 0.686 | 17.663 ± 0.967 | 17.378 ± 0.993 | 16.167 ± 1.189 | 15.746 ± 1.016 | 14.952 ± 1.127 | <0.001 |
| *Vessel Density* | 37.163 ± 1.115 | 36.491 ± 1.382 | 36.278 ± 1.469 | 34.648 ± 1.881 | 34.592 ± 1.854 | 33.052 ± 1.838 | <0.001 |

\* NDR: no diabetic retinopathy
NPDR: non-proliferative diabetic retinopathy
PDR: proliferative diabetic retinopathy
FAZ: foveal avascular zone

Linear regression analyses were performed to quantify the amount of change in each microvasculature index over time, and the slopes of the regression lines were used to compare the relative change in each parameter between DR categories (Figure 4, 5). In the deep layer, intergroup comparison found a significant decrease in skeleton density (-0.56 ± 0.39% per year, p = 0.04) and vessel density (-0.13 ± 0.29% per year, p = 0.01) over time in patients with severe NPDR compared to healthy controls. Additionally, skeleton density showed a significant increase in patients with PDR (0.10 ± 0.84% per year, p = 0.04) compared to patients with severe NPDR. Acircularity index in the deep layer increased significantly in patients with severe NPDR (0.16 ± 0.12% per year, p = 0.05) compared to those with mild NPDR. In the superficial layer, the average loss of vessel caliber in patients with severe NPDR (-4.5e-4 ± 0.0005% per year, p = 0.04) was significantly greater than that of patients with mild NPDR, who on average gained 3.15e-2 microns in vessel caliber per year (Figure 5, Table 3).

**Figure 4.**
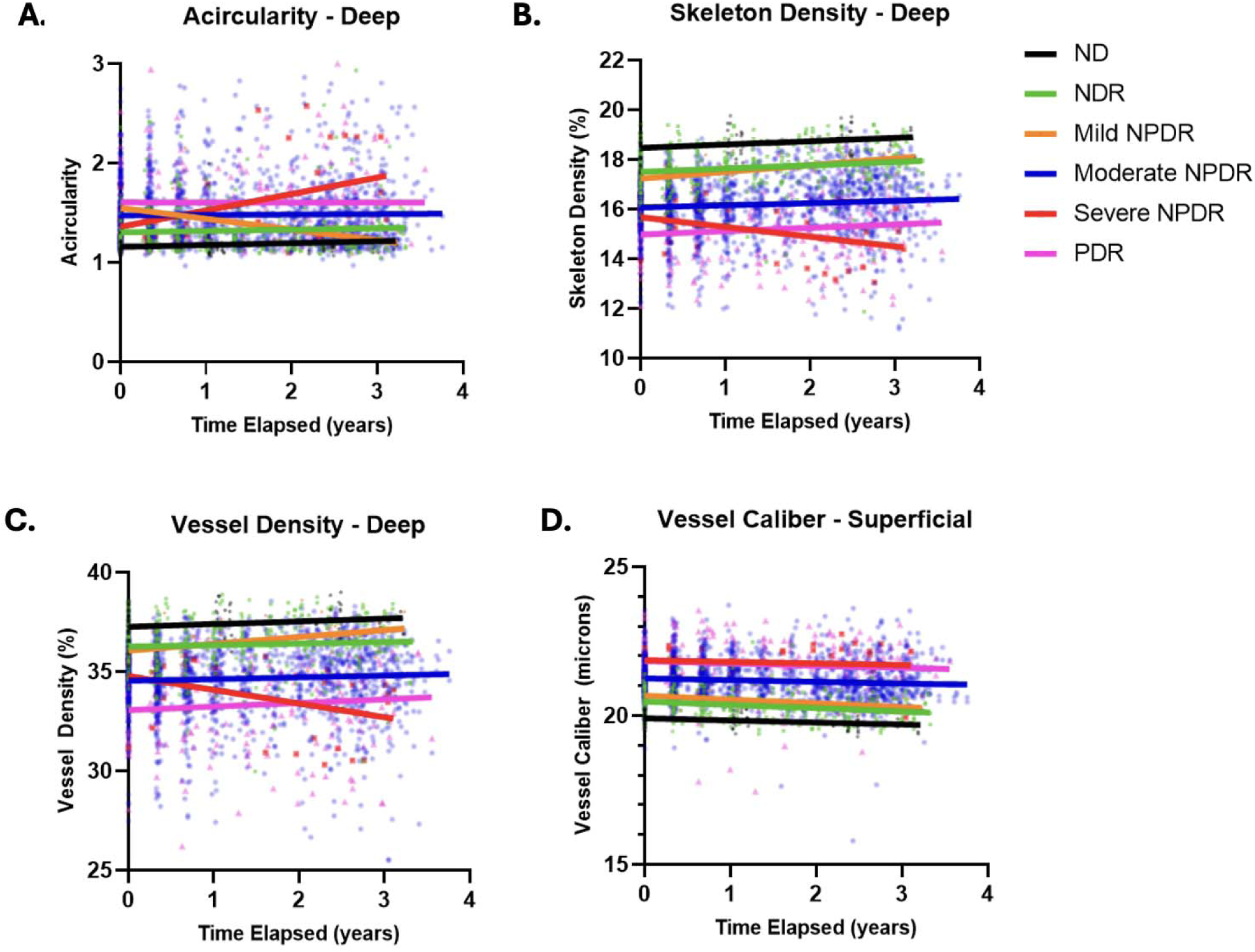
Change in microvasculature indices over time. DR classes are as follows: Non-diabetics (ND}, diabetics with no diabetic retinopathy (NDR), Mild Non-Proliferative Diabetic Retinopathy (Mild NPDR), Moderate Non­ Proliferative DR (Moderate NPDR}, Severe Non-Proliferative DR (Severe NPDR}, and Proliferative DR (PDR}. Each data point represents an individual patient measurement, and the solid lines represent best-fit linear regression lines for each DR class.

**Figure 5.**
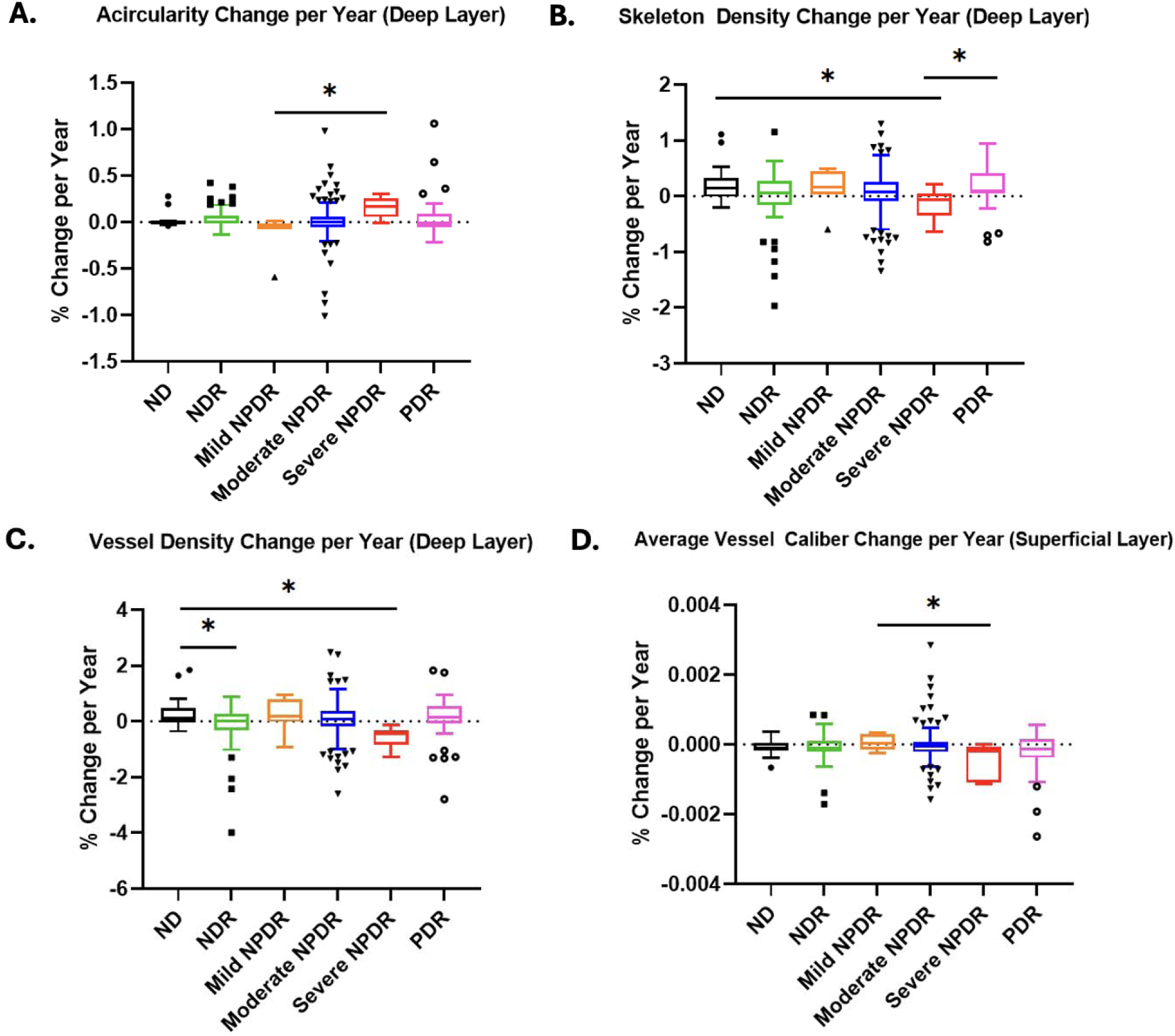
Average yearly change in microvasculature indices of vessel density, skeleton density, acircularity, and average vessel caliber differ significantly over time across DR classes. Boxplots show median and IQR. ND = non-diabetics, NDR = no diabetic retinopathy, NPDR = non-proliferative diabetic retinopathy, PDR = proliferative diabetic retinopathy. Asterisks indicate significant differences(* p < 0.05)

**Table 3:** Vessel metrics percent change per year between DR classes. Data presented as mean ± standard deviation (% change per year).

|  | Healthy controls | NDR | Mild NPDR | Moderate NPDR | Severe NPDR | PDR | *p* value |
| --- | --- | --- | --- | --- | --- | --- | --- |
| **<u>Deep layer:</u>** |  |  |  |  |  |  |  |
| Acircularity | 0.011±0.07 | 0.035±0.11 | -0.12±0.21 | 0.007±0.18 | 0.16±0.12 | 0.05±0.24 | **0.045 \*** |
| Average Vessel Caliber | -7.1e-2±0.12 | -4.9e-2±0.27 | -2.7e-2±0.15 | -3.8e-2±0.28 | -0.15±0.25 | -0.18±0.57 | 0.180 |
| FAZ Area | -0.006±0.012 | -0.012±0.100 | -0.047±0.141 | 0.002±0.064 | 0.001±0.056 | 0.021±0.122 | 0.219 |
| FAZ Perimeter | -0.023±0.058 | -0.034±0.603 | -0.528±1.442 | 0.025±0.615 | 0.336±0.550 | 0.239±0.961 | 0.058 |
| Fractal Dimension | 3.4e-4±0.0008 | -5.7e-4±0.003 | 6.9e4±0.001 | -6.38e-5±0.003 | -5.1e-4±0.002 | 7.7e-4±0.003 | 0.723 |
| Skeleton Density | 0.203±0.310 | -0.025±0.509 | 0.143±0.362 | 0.066±0.355 | -0.296±0.232 | 0.174±0.371 | **0.010 \*** |
| Vessel Density | 0.276±0.502 | -0.151±0.810 | 0.252±0.624 | 0.066±0.583 | -0.563±0.391 | 0.097±0.839 | **0.011 \*** |
| **<u>Superficial Layer:</u>** |  |  |  |  |  |  |  |
| Acircularity | -0.007±0.0346 | 0.035±0.191 | -0.007±0.157 | 0.018±0.157 | -0.147±0.126 | 0.003±0.208 | 0.174 |
| Average Vessel Caliber | -8.6e2± 0.02 | -9.1e-2 ±0.04 | 3.1e-2 ±0.02 | -3.6e-2±0.04 | -4.5e-1 ±0.05 | -2.3e-1±0.06 | **0.0396 \*** |
| FAZ Area | -0.002±0.009 | -0.010±0.076 | -0.002±0.010 | 0.00240.086 | -0.091±0.068 | 0.006±0.119 | 0.175 |
| FAZ Perimeter | -0.025±0.098 | 0.001±0.398 | -0.030±0.141 | 0.060±0.790 | -0.905±0.742 | 0.036±1.160 | 0.083 |
| Fractal Dimensions | 3.4e-4±0.0008 | -5.7e-4±0.003 | 6.9e4±0.001 | -6.38e-5±0.003 | -5.1e-4±0.002 | 7.7e-4±0.003 | 0.444 |
| Skeleton Density | 0.161±0.258 | 0.064±0.358 | 0.073±0.302 | 0.063±0.533 | 0.398±0.775 | 0.226±0.751 | 0.325 |
| Vessel Density | 0.223±0.357 | 0.011±0.408 | 0.206±0.505 | 0.109±0.794 | 0.256±1.127 | 0.198±1.197 | 0.812 |

\* NDR: no diabetic retinopathy
NPDR: non-proliferative diabetic retinopathy
PDR: proliferative diabetic retinopathy
FAZ: foveal avascular zone

## Discussion

Our study provides important insights into the longitudinal changes in retinal microvascular indices associated with DR. Our 3-year follow up data reveals significant rates of change in vessel density, skeleton density, acircularity index, and average vessel caliber between DR severity groups. These results confirm the utility of OCTA in detecting microvascular changes associated with DR [16, 17] and adds to our current knowledge by demonstrating rates of change of these OCTA metrics over 3 years.

One of the key findings of our study is the progressive decline in vessel density and skeleton density in the deep capillary plexus in patients with severe NPDR. Patients with severe NPDR showed a significant annual decrease in skeleton density and vessel density compared to healthy controls who demonstrated annual increases in these metrics. This accelerated capillary dropout likely reflects advancing ischemia and corresponds with the pathophysiological transition from moderate to severe disease where widespread capillary non-perfusion becomes evident. This reduction in capillary perfusion aligns with prior studies that have identified vessel density as a reliable anatomic marker of DR severity [18, 19]. The observed difference in rates of vessel density loss between NPDR stages suggest that these changes may serve as early indicators of subtle disease progression, which has significant clinical implications for monitoring at-risk patients. Surprisingly, our study also found an increase in skeleton density in patients with PDR compared to severe NPDR, which may reflect compensatory revascularization or onset of neovascularization, a hallmark of advanced DR [20].

Significant changes in the FAZ, as measured by the acircularity index, provide insight into the progression of macular ischemia in DR and DME. Acircularity increased significantly in severe NPDR compared to mild NPDR, suggesting worsening ischemic damage as DR progresses leading to permanent visual impairment. This progressive irregularity in FAZ morphology may represent uneven capillary dropout along the FAZ margin, creating a more tortuous and irregular border as the disease advances. This observation is consistent with findings from longitudinal OCTA studies that report FAZ enlargement and irregularity as indicators of worsening microvascular dysfunction in DR [21, 22]. The increasing acircularity without necessarily showing significant changes in FAZ area in the same timeframe suggests that border irregularity may be a more sensitive anatomic marker of early ischemic damage than simple FAZ enlargement. Clinically, this may explain why patients with DME do not regain full visual potential despite resolution of DME with anti-VEGF injections [23].

The observed reduction in average vessel caliber in the superficial capillary plexus in severe NPDR suggests progressive capillary narrowing, which may contribute to impaired blood flow and oxygen delivery to the inner retina. While some prior studies have shown a positive association between vessel caliber and DR severity, microvascular constriction often precedes capillary dropout in DR and may be an early sign of disease progression [24–26]. The significant differences in vessel caliber changes between mild and severe NPDR in our study highlights its potential as an early anatomic marker of disease severity.

Our findings have important clinical implications. First, the identification of specific longitudinal markers, particularly deep plexus vessel and skeleton density, acircularity index, and superficial plexus vessel caliber, suggest that OCTA can be useful for monitoring of DR progression. Based on our findings, we propose that a >0.5% annual decrease in deep plexus skeleton density or a vessel caliber reduction exceeding 0.3 micrometers per year may signal high-risk disease. This may allow for earlier detection of high-risk patients and more timely intervention, potentially reducing the risk of vision-threatening complications. Moreover, the observed variability in OCTA metrics across different DR stages suggests that individualized monitoring strategies based on disease severity may be beneficial, with patients showing early changes in these metrics potentially requiring more frequent assessment than those with stable measurements. This would allow more optimal allocation of increasingly limited healthcare resources. Finally, our findings suggest that layer-specific analysis is essential, as the deep capillary plexus showed more consistent changes associated with disease progression than the superficial plexus, which aligns with the understanding that DR often affects deeper vessels first. Future studies should focus on standardizing OCTA acquisition protocols and further investigating the relationship between these microvascular indices and functional outcomes such as visual acuity.

Several limitations of our study must be acknowledged. The relatively small subset sample size, particularly in the severe NPDR group (n = 6), may limit the generalizability of our findings and requires validation in larger cohorts. Additionally, while we controlled for image quality by excluding images with signal strength ≤ 5, other potential confounding factors such as glycemic control fluctuations, blood pressure variation and medication changes during the follow-up period could influence microvascular metrics independent of DR progression. Our imaging processing pipeline, while systematic, may introduce measurement variability, particularly for smaller vessels near the resolution limit of the OCTA technology we used. Missing data points may have influenced our ability to detect subtle changes in OCTA metrics, particularly in the PDR group where clinical interventions such as anti-VEGF injections and peripheral laser treatments could have altered the natural disease course.

In conclusion, our 3-year longitudinal study provides novel insight into the dynamic changes of retinal microvasculature across the spectrum of DR severity. By identifying specific OCTA biomarkers that correlate with disease progression, we provide a foundation for improved DR monitoring and early intervention strategies using a safe, non-invasive, and rapid imaging modality. Future research should focus on validating these anatomic markers in larger cohorts, establishing normative databases of progression rates, and determining whether interventions targeted at patients showing accelerated changes in these metrics can prevent vision-threatening complications. Continued research and longitudinal studies are essential to further validate these anatomic biomarkers and establish standardized protocols that can be integrated into clinical practice.

## Data Availability

All data produced in the present study are available upon reasonable request to the authors.

